# On the limitations of large language models in clinical diagnosis

**DOI:** 10.1101/2023.07.13.23292613

**Authors:** Justin T Reese, Daniel Danis, J Harry Caufield, Tudor Groza, Elena Casiraghi, Giorgio Valentini, Christopher J Mungall, Peter N Robinson

**Affiliations:** Division of Environmental Genomics and Systems Biology, Lawrence Berkeley National Laboratory, Berkeley, CA, 94720, USA; The Jackson Laboratory for Genomic Medicine, Farmington CT, 06032, USA; Rare Care Centre, Perth Children’s Hospital, Perth, WA 6009, Australia; Telethon Kids Institute, Perth, WA 6009, Australia; AnacletoLab, Dipartimento di Informatica, Università degli Studi di Milano, Milano, Italy; ELLIS-European Laboratory for Learning and Intelligent Systems; Institute for Systems Genomics, University of Connecticut, Farmington, CT 06032, USA

**Keywords:** Large language model, Generative Pretrained Transformer 4, Artificial Intelligence, Differential diagnosis

## Abstract

**Objective:** Large Language Models such as GPT-4 previously have been applied to differential diagnostic challenges based on published case reports. Published case reports have a sophisticated narrative style that is not readily available from typical electronic health records (EHR). Furthermore, even if such a narrative were available in EHRs, privacy requirements would preclude sending it outside the hospital firewall. We therefore tested a method for parsing clinical texts to extract ontology terms and programmatically generating prompts that by design are free of protected health information.

**Materials and Methods:** We investigated different methods to prepare prompts from 75 recently published case reports. We transformed the original narratives by extracting structured terms representing phenotypic abnormalities, comorbidities, treatments, and laboratory tests and creating prompts programmatically.

**Results:** Performance of all of these approaches was modest, with the correct diagnosis ranked first in only 5.3-17.6% of cases. The performance of the prompts created from structured data was substantially worse than that of the original narrative texts, even if additional information was added following manual review of term extraction. Moreover, different versions of GPT-4 demonstrated substantially different performance on this task.

**Discussion:** The sensitivity of the performance to the form of the prompt and the instability of results over two GPT-4 versions represent important current limitations to the use of GPT-4 to support diagnosis in real-life clinical settings.

**Conclusion:** Research is needed to identify the best methods for creating prompts from typically available clinical data to support differential diagnostics.

## INTRODUCTION

Large language models (LLMs) are a class of artificial intelligence (AI) algorithms that are trained on billions of words of diverse texts using self-supervised learning techniques.^1^ The resulting models can be used for natural language processing (NLP) tasks such as chatbots and text prediction.^2^ LLMs are designed for one-shot or few-shot learning, meaning that they can perform well on a wide range of tasks without specific training, or can be pre-trained on huge corpora of text and then can be secondarily fine-tuned on specific tasks using relatively small training datasets.^3^ Most previous machine learning (ML) algorithms were trained to only perform specific tasks, and in contrast to LLMs are usually not able to address previously unseen problems without needing to be completely retrained.^4^

ChatGPT is a chatbot that can be “prompted” to perform a range of tasks; examples of the task to be performed can be provided in the input to improve performance. ChatGPT uses the Generative Pretrained Transformer (GPT) as its backend LLM. In contrast to some previous LLMs that were trained to predict the next word in web pages, GPT-4 uses reinforcement learning from human feedback to fine-tune the model to follow a broad class of written instructions.^5^ LLMs have achieved competitive performance in answering questions derived from or similar to Medical Licensing Examination questions,^6,7^ and ChatGPT has been shown to provide useful answers to patients’ queries.^8^ Several studies have been published on the performance of LLMs in generating a differential diagnosis in response to a query that summarizes the presenting signs and symptoms.^9–13^ The methods and assessment approaches differed between the studies, but in general, the studies constructed a prompt asking for the differential diagnosis for a patient followed by a clinical case vignette as full text. The performance in these studies was deemed by the authors to be impressive, but currently only suitable for supporting physician judgment rather than replacing it.

A recent study^14^ reported that the Generative Pretrained Transformer 4 (GPT-4) model performed well in complex differential diagnostic reasoning. They evaluated the performance of GPT-4 on 75 case records from the New England Journal of Medicine (NEJM) published in 2021 and 2022 by giving GPT-4 a standard prompt, followed by the part of the case report that included the case presentation up to but not including the discussant’s initial response. The authors found that GPT-4 returned the correct diagnosis as a part of its response in 64% of cases, with the correct diagnosis being at rank 1 in 39% of cases.^14^

## OBJECTIVE

We examined the influence of linguistic context on the performance of GPT-4 in differential diagnostic tasks by comparing prompts characterized by a rich narrative context, as those that contained the clinical data described in the 75 NEJM case reports, with more schematic prompts which more closely mimic the clinical data effectively available in clinical practice.. More precisely, we compared three different approaches to generating prompts and two versions of GPT-4 to determine the stability of the performance of GPT-4 in differential diagnostic challenges.

## MATERIALS AND METHODS

We included 75 NEJM case reports from Case 2-2021 to 40-2022 in our study, omitting 5 of the 80 case reports that did not describe a diagnostic dilemma.^14^ The text representing the initial clinical presentation was taken from the documents by extracting the first discussant’s section. We appended this text to the standardized prompt as described in the recent study,^14^ and used the OntoGPT^15^ tool to query GPT-4. To assess the influence of the narrative context of the case reports, we extracted the clinical abnormalities as Human Phenotype Ontology (HPO) terms.^16^ Observed and excluded abnormalities were included in the prompt using a standard template, whereby the clinical features were arranged according to the time point of presentation (Figure 1).

**Figure 1.**
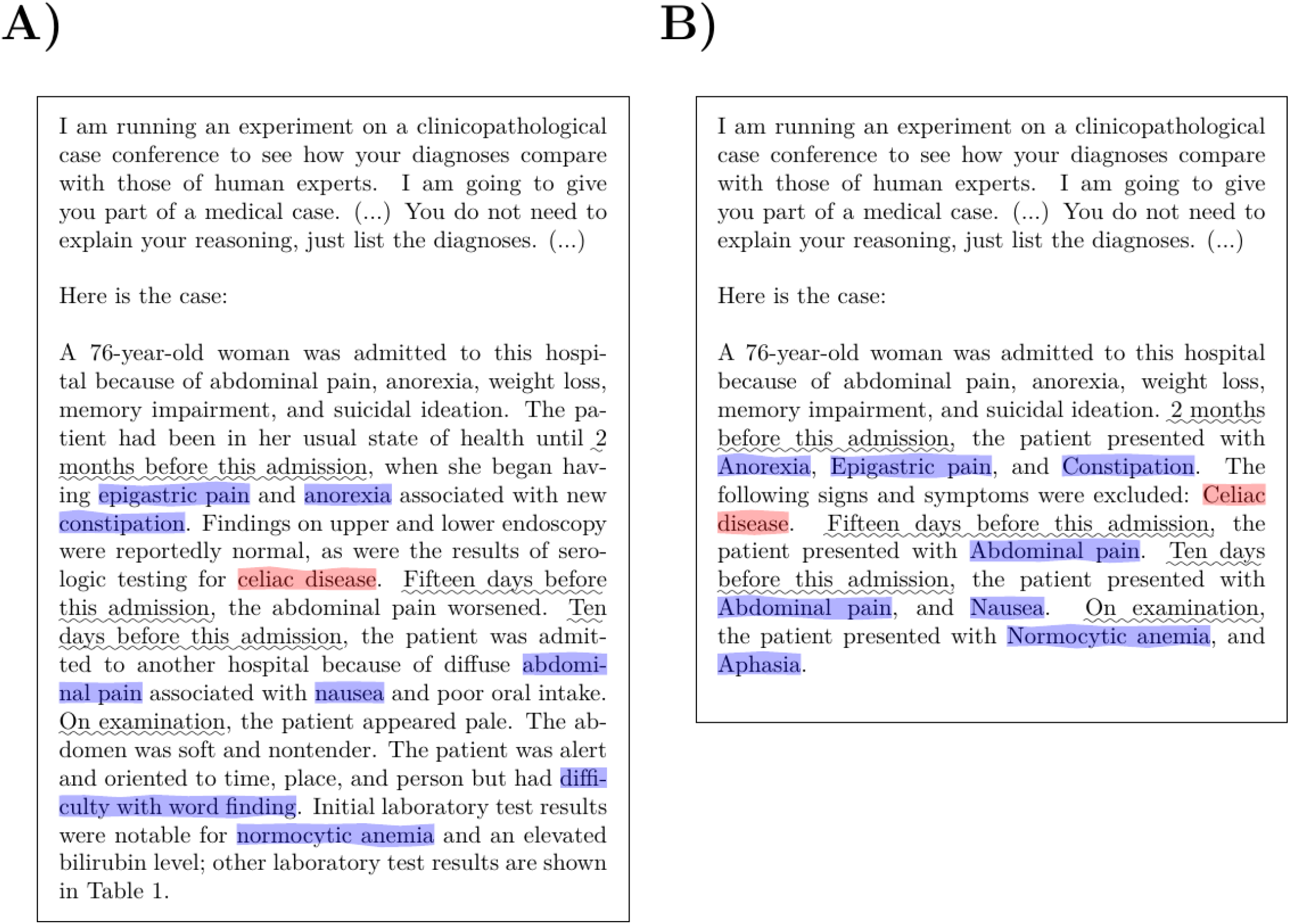
GPT-4 prompt templates. (A) An example showing the narrative prompt provided to GPT-4 (case 38-2021). (B) The corresponding example using the programmatically generated prompt (PHENO-R). The features in the original text that were mined to generate the feature-based query are highlighted (observed features in blue and excluded features in red; the phrases describing the time periods are underlined). In this example, the case presentation is relatively short; in many of the analyzed cases, the presentation had a length of a page or longer. The full preamble text for this example is given in Supplemental Figure S1.

### Prompt generation approaches

The text that is sent as input to the GPT-4 models is referred to as a prompt. For the experiments reported here, we created prompts with a preamble that instructs GPT-4 on the desired response and a second part with information about the clinical case. We used the preamble text as previously reported for a study that assessed the performance of GPT-4 with diagnostic challenges.^14^ The preamble begins with the sentences “I am running an experiment on a clinicopathological case conference to see how your diagnoses compare with those of human experts. I am going to give you part of a medical case.” and includes the instructions “the goal is to get the correct answer, not a broad category of answers. You do not need to explain your reasoning, just give the diagnosis/diagnoses.” We tested four methods to generate the portion of the prompt after the preamble that contained information about the clinical case in question. Each method used the same preamble followed by text that described the clinical case.

#### 1. Narrative prompt generation (NARR)

Here, the original text of the NEJM case reports including only the section of the first discussant (i.e., the physician who presents the first part of the clinical history). For instance, Case 26-2022 begins with this: “Dr. Esra D. Gumuser (Medicine): A 48-year-old woman was admitted to this hospital because of multiple lung and liver lesions identified during an evaluation for abdominal pain….” and continues with a discussion of the initial clinical presentation.^17^ An accurate prediction of the correct diagnosis would have the highest value at this initial point in the clinical course, because it could guide confirmatory diagnostics and subsequent clinical management. The NEJM case reports continue with contributions of other discussants (i.e., other physicians who were involved with the case) about further diagnostics or considerations of specific differential diagnoses. For instance, in our example, the case report continues with “Dr. Melissa C. Price: A chest radiograph showed faint nodular opacities in the lungs, predominantly in the upper lobes….”. Sections such as this subsequent to the first section of the first discussant were not included in the prompt. An example is shown in Supplemental Figure S2.

#### 2. Phenotypic feature prompt generation (PHENO-R and PHENO-C)

Here, the narrative text was subjected to concept recognition to identify Human Phenotype Ontology (HPO) terms using the fenominal text mining tool (https://github.com/monarch-initiative/fenominal). The HPO is a comprehensive resource that systematically defines and logically organizes human phenotypes. Broad clinical, translational and research applications using the HPO include genomic interpretation for diagnostics, gene-disease discovery, mechanism discovery and cohort analytics, all of which assist in facilitating precision medicine.^16,18,19^ Regular expressions were used to identify phrases that marked separate presentations at specific ages or in specific situations, such as “3 days before this admission” and “After 3 weeks of treatment”. These were used to segment the original narrative into sections, and then a template was used to create text describing the clinical manifestations. The fenominal tool classifies matches as observed or excluded, and the template was designed accordingly.

For instance, consider a text segment that contains the phrase “Three months before admission”. If text mining by fenominal identified the observed HPO terms Splenomegaly (HP:0001744) and Cholelithiasis (HP:0001081) and the excluded (negated) HPO terms Dyspnea (HP:0002094) and Orthopnea (HP:0012764), the template would output the following:

> *Three months before admission, the patient presented with Splenomegaly and Cholelithiasis. The following signs and symptoms were excluded: Dyspnea and Orthopnea*.

Two versions of phenotypic feature prompt generation were assessed. In the first, the order of the time periods was random (PHENO-R), and in the second, the order of the time periods was chronological (PHENO-C). An example of each is shown in Supplemental Figure S3 and S4. Figure 1 shows a comparison of the NARR and PHENO-R prompt generation approaches.

#### 3. Manual/HPO prompt generation (MAN-HPO)

Here, manual curation of medical history, family history, and current treatments was used to supplement the PHENO-C approach. The template for the PHENO-C approach was extended accordingly. An example is shown in Supplemental Figure S5.

Prompts and GPT-4 output for all 75 case reports for each of the four approaches are available in the zenodo repository:

https://zenodo.org/record/8353301

We developed a Java application for generation of prompts as described above called phenopacket2prompt that is available under an open-source MIT license at https://github.com/monarch-initiative/phenopacket2prompt. Version 0.3.7 was used for the analysis described here and is available as a tagged release in the GitHub repository.

### Assessment of GPT4 responses

GPT-4 was queried by sending prompts to the OpenAI API using OntoGPT (https://github.com/monarch-initiative/ontogpt), a tool that includes a programmatic interface to the Open AI API. The response from GPT-4 for each prompt was saved in a TSV file. The differential diagnoses returned by GPT-4 were then transferred to spreadsheets and independently assessed by J.T.R. and P.N.R. Disagreements were resolved by discussion. The results were assigned scores using the same scale as in the previous study.^14^ Results were scored independently by three coauthors (P.N.R, D.D., J.T.R.) and disagreements were resolved by consensus. 5 = the actual diagnosis was suggested in the differential; 4 = the suggestions included something very close, but not exact; 3 = the suggestions included something closely related that might have been helpful; 2 = the suggestions included something related, but unlikely to be helpful; 0 = no suggestions close to the target diagnosis.

## RESULTS

Several recent studies have explored the utility of LLMs including GPT in supporting differential diagnosis.^9–14^ The prompts that are submitted to the LLMs consist of a preamble with instructions about the desired response and a clinical vignette containing information about the case. In one study, clinical vignettes were taken verbatim from the case studies published in the New England Journal of Medicine (NEJM).^14^

We posited that the clinical vignettes available in the NEJM case reports are not representative of texts that could actually be submitted to LLM decision support tools. Although it is difficult to quantify, the texts are written in an almost literary style and present detailed and comprehensive clinical information in a logical sequence. We estimate that it would take many hours if not days to create such a vignette from typical clinical notes, laboratory results, and imaging findings. Even if it were possible to automatically generate a clinical vignette of comparable quality directly from electronic health records (EHRs), privacy regulations would prohibit these vignettes from being transmitted to external LLMs such as chatGPT because of the possibility that the vignettes contain protected health information (PHI).

One way to use LLMs in clinical routine that would avoid these difficulties would be to automatically extract the salient information about a case in structured form using standard terminologies that by design cannot contain PHI, and then to programmatically generate a query based on the structured information. Here, we explore the performance of GPT-4 as a differential diagnostic tool using the original narratives from the NEJM case reports to generate prompts and three methods for generating prompts on the basis of structured data, or structured data supplemented by manual curation.

The approaches to prompt generation are denoted by NARR, PHENO-R, PHENO-C, and MAN-HPO, and are explained in the Methods.

### Narrative and feature-based prompts

We presented GPT-4 with the original description by the primary discussant (NARR approach), and observed that GPT-4 included the final correct diagnosis in its differential in 29/75 (38.7%) of cases, and the correct diagnosis was in the first position (rank 1 and score of 5) in 17.3% of cases. The mean rank for any helpful diagnosis (with a score of at least 3) was 3.4. These results are similar to but not identical to those of a study by Kanjee et al. on the same input data.^14^ We then tested the PHENO-R approach, which generates a simple narrative text based on a structured representation of phenotypic features as HPO terms (Methods). Here, GPT-4 included the final diagnosis in its differential in only 8/75 (10.7%) of cases, and the correct diagnosis was in the first position (rank 1 and score of 5) in 4.0% of cases. The mean rank for any helpful diagnosis was 3.9. (Figure 2).

**Figure 2.**
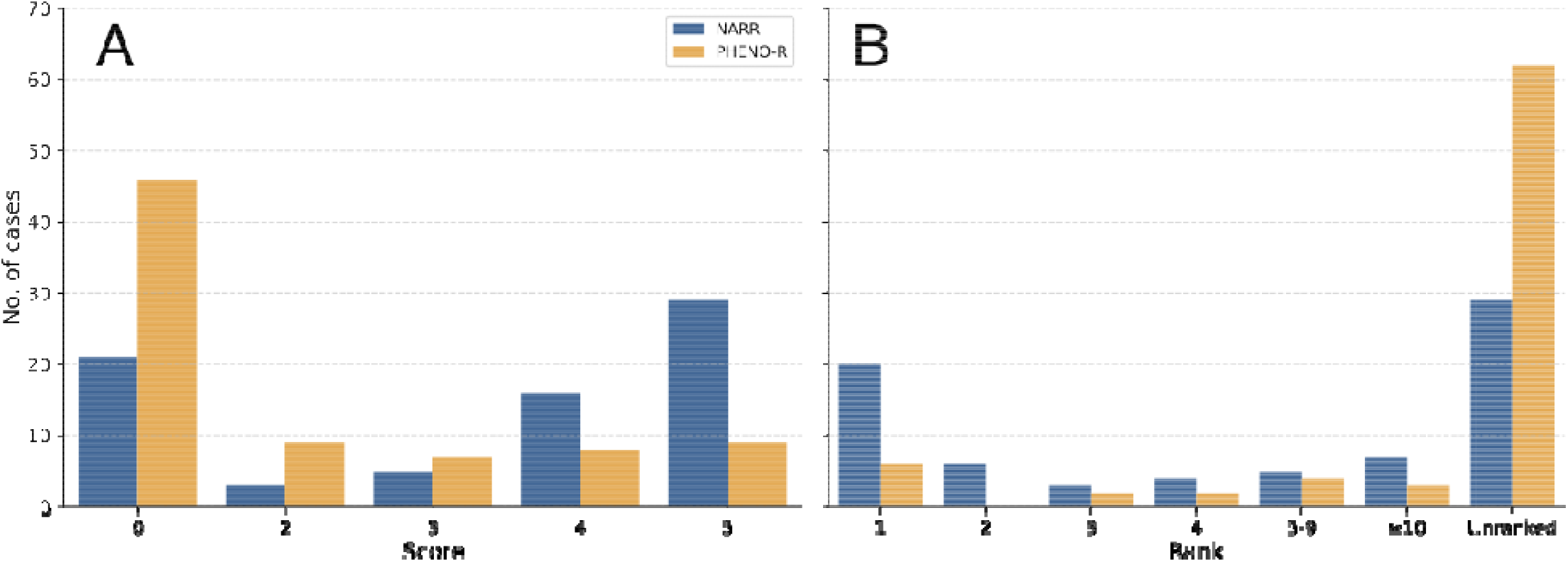
Performance of GPT-4 in diagnostic challenges using narrative (NARR) or structured (phenotypic feature-PHENO-R) prompts. (A) A histogram of scores (0-5) denoting accuracy of GPT-4 diagnosis using narrative (blue bars) and extracted features (orange bars) from NEJM case reports. (B) A histogram of the ranks of the correct diagnosis in the differential diagnosis produced by GPT-4 in cases where the score was 4 (nearly the correct diagnosis) or 5 (correct diagnosis) using NARR (blue bars) and PHENO-R (orange bars) from NEJM case reports. The count of unranked cases (scores 0-3) are shown for comparison. The results were assigned scores using a previously published scale (See Methods for details).

**Figure 3.**
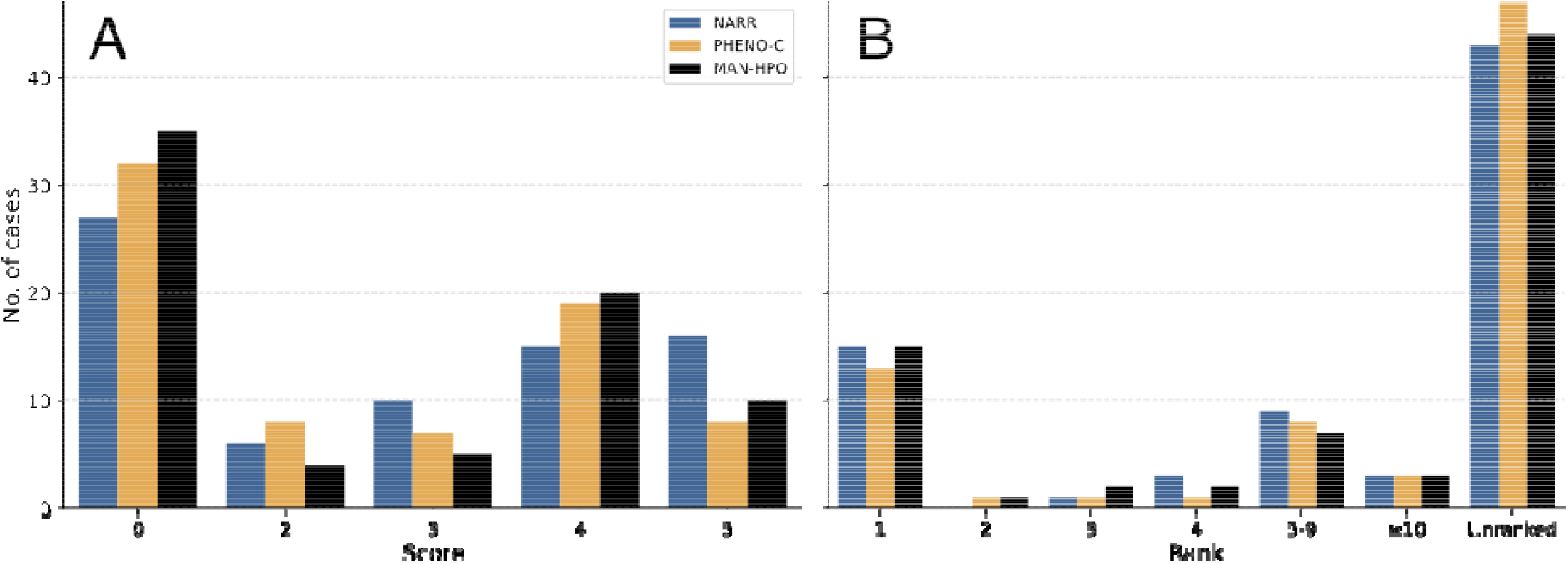
Performance of GPT-4 in diagnostic challenges using narrative, feature-based, or manually annotated prompts. Histogram of scores (0-5) denoting accuracy of GPT-4 diagnosis using narrative (blue bars), extracted features (orange bars), or manual annotation (black bars) from NEJM case reports. (B) A histogram of the ranks of the correct diagnosis in the differential diagnosis produced by GPT-4 in cases where the score was 4 (nearly the correct diagnosis) or 5 (correct diagnosis) using narrative (blue bars), extracted features (orange bars), or manual annotation (black bars) from NEJM case reports. The count of unranked cases (scores 0-3) are shown for comparison. The results were assigned scores as in Figure 2.

**Figure 4.**
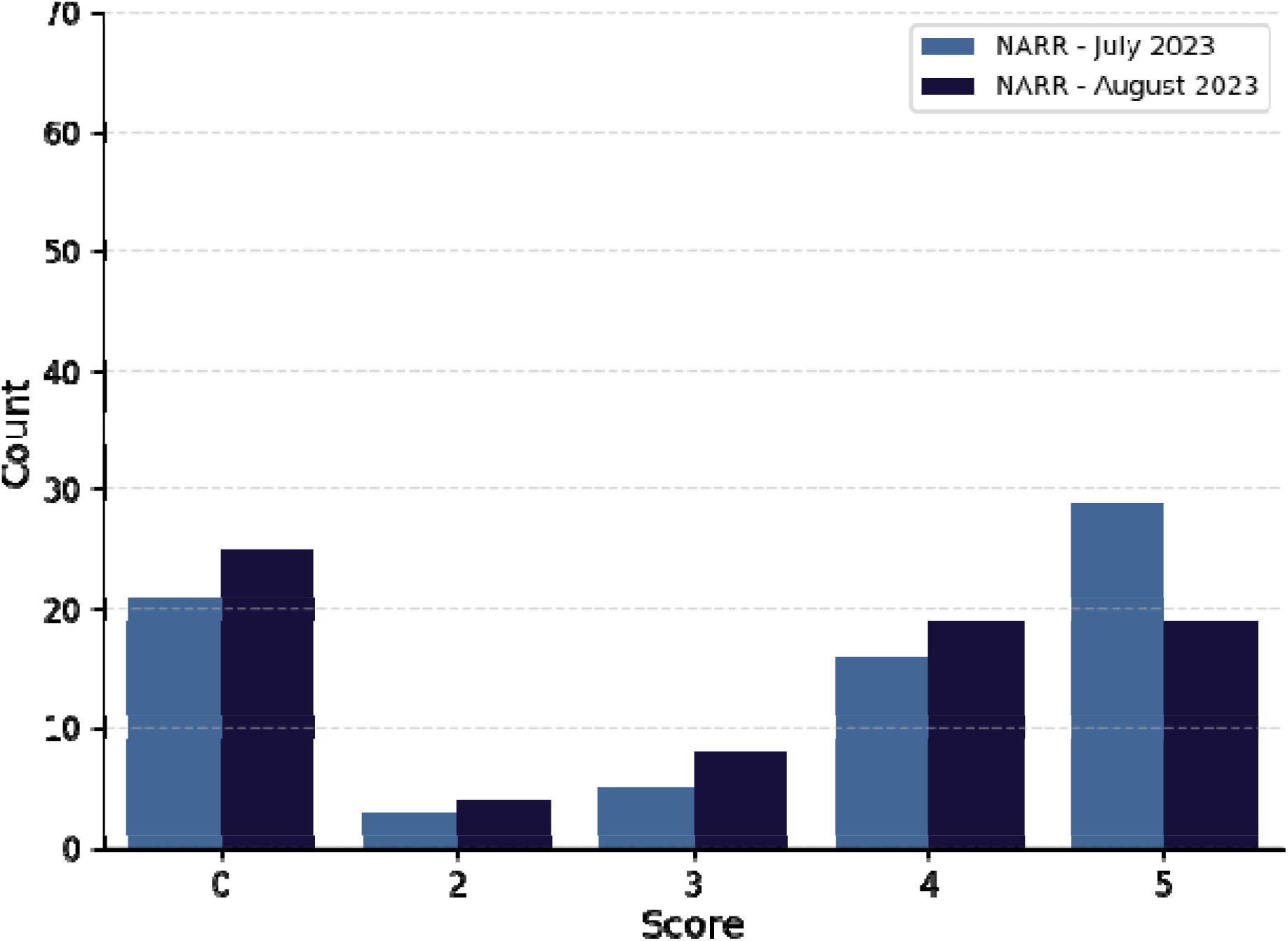
Performance of GPT-4 in diagnostic challenges using narrative prompts in July 2023 and August 2023. Narrative prompts for each of the 75 cases were prepared as described previously. A histogram of scores (0-5) denoting accuracy of GPT-4 diagnosis in July 5, 2023 (light blue bars, left) and August 25, 2023 (dark blue bars, right) from NEJM case reports.

### Manual prompt curation

The fact that the results of the phenotypic feature-based prompt were inferior to those of the narrative prompt suggested that either the narrative style of the original text was important for the performance of GPT, or that essential information had been lost by our feature extraction process, which exclusively extracts clinical phenotypes encoded by HPO terms. Therefore, we manually curated the 75 case reports to add information about past medical history, family history, and current treatments. While the experiments described in the previous section were performed on July 5, 2023 (version gpt-4-0314), the following experiments were carried out on August 29, 2023 (GPT-4 version gpt-4-0613). The GPT-4 API allows use only of the most current model version.

We presented GPT-4 with narrative (NARR), phenotypic feature-based (PHENO-C; see Methods), and manually annotated (MAN-HPO) prompts. We observed that using NARR, GPT-4 included the correct diagnosis in its differential in 16/75 (21.3%) of cases, and the correct diagnosis was in the first position (rank 1 and score of 5) in 10.6% of cases. The mean rank for any helpful diagnosis (score of >=3) was 4.1). One of the cases failed to be processed by GPT-4 and was assigned the score of 0. Using the PHENO-C prompts, GPT-4 included the correct diagnosis in 8/75 (10.7%) of cases, and the correct diagnosis was in the first position (rank 1 and score of 5) in 5.3% of cases. The mean rank for any helpful diagnosis (score of >=3) was 4.6. Using manually annotated (MAN-HPO) prompts, GPT-4 included the correct diagnosis in 10/75 (13.3%) of cases, and the correct diagnosis was in the first position (rank 1 and score of 5) in 10.6% of cases. The mean rank for any helpful diagnosis (score of >=3) was 4.2.

### Stability of results across GPT4 versions

We noted that in both of the above analyses our results with the narrative-queries were less good than those reported by Kanjee et al. on the same dataset.^14^ We therefore repeated the analysis originally performed on July 5, 2023 with the version of GPT4 current at the time of this writing (August 30, 2023;) on the narrative prompts.

Scant details are available about differences between different versions of GPT with respect to algorithmic details and training data.^20^ We compared the results of our experiment using the text-only query corpus obtained on July 5 2023 (version gpt-4-0314) and August 29, 2023 (GPT4 version gpt-4-0613). In July, there were 29 correct diagnoses (score of 5, 38.6%) as compared to only 19 (25.3%) for the August version; on the other hand, 21 (28%) queries returned no relevant information in the July version (score of 0), compared to 25 (33.3%) for the August version.

## DISCUSSION

The potential of large language models (LLM) such as GPT has been a subject of debate, with some ascribing near sentient abilities to the models and others claiming that LLMs merely perform “autocomplete on steroids.” For the purpose of applying LLMs to the problem of clinical diagnosis, it is important to realize that LLMs generate text based on patterns learned from huge amounts of training texts.^21^ LLMs such as GPT-4 do not possess an explicit model of medical domain knowledge and do not perform a symbolic human-like reasoning, but instead perform autocompletion by implicitly learning medical domain knowledge from the data.

One hypothesis we explore in this study is that the narrative form in which information is presented to GPT-4 influences the performance of GPT-4 in differential diagnostic tasks. We first compared the performance of GPT-4 on the original narrative texts and simplified versions of the cases in which only clinical features representable by HPO terms are presented to GPT-4. The performance on the feature-based queries was substantially worse than that of the narrative queries (Figure 2). We reasoned that our method of identifying phenotypic abnormalities from the clinical vignette by recognition of HPO terms did not identify all relevant information. Therefore, we manually curated the past medical history, family history, comorbidities, abnormal laboratory findings that had been missed by the HPO mining, and treatments. While this approach improved the performance, it was still substantially worse than that using the original texts. Additionally, we noted that the performance of GPT-4 on the identical prompts was substantially different for two different versions of GPT-4 (gpt-4-0613 and gpt-4-0314).

We consider the feature-based queries to be a more appropriate test of the performance of GPT-4 in diagnostic tasks, since it is unlikely that the narrative approach can be used in actual clinical practice. NEJM-style clinical narratives are not readily available for most cases and EHR texts cannot be transmitted across the internet without violating privacy regulations. In contrast, it is straightforward to generate a feature-based list of clinical problems, symptoms, and other abnormalities that can be used to generate a prompt for GPT. Currently, GPT-4 is not available for installation within medical centers, and it remains an open question as to whether smaller models, eventually embedding structured information, will demonstrate comparable performance. A possible solution could consist in coupling LLMs with a formal representation of medical knowledge, for example using biomedical knowledge graphs.^4^

### Strengths and Limitations

We did not systematically test the variation in responses of GPT-4 related to the stochasticity of the algorithm, which should be the subject of future work. To rigorously perform such a study, the community would need to develop a framework for measuring variability in a standardized fashion that would determine how to deal with various degrees of textual differences in the responses (e.g., pregnancy associated cardiomyopathy vs. cardiomyopathy during pregnancy), how to grade matches of general and specific diagnoses (e.g., Idiopathic multicentric Castleman’s disease vs. Castleman’s disease), how to assess failure of GPT-4 to provide a genetic diagnosis (e.g., the correct diagnosis is “Relapsed acute myeloid leukemia (with wild-type NPM1 and newly identified internal tandem duplication mutation in FLT3” and GPT-4’s diagnosis is simply “Relapsed acute myeloid leukemia”). The assessment scale we used in this study measures how often the correct diagnosis was at any rank in the list returned by GPT-4, but clearly the utility of the prediction is lower if the diagnosis is at the low range of a long list.

Our study did not comprehensively explore all possible ways of generating prompts from structured data. However, it does point out two important limitations of the LLMs for clinical diagnostics that will need to be addressed in order to apply the method to support differential diagnosis in routine clinical care. Firstly, we show that the way the prompt is generated can have substantial effects on performance. Although the full narrative text had the best performance, it would be challenging to create a similar narrative text from data available in typical EHRs or other clinical settings. Therefore, we contend that approaches that extract features automatically from available clinical texts, or that derive ontology terms from structured data such as laboratory values,^22^ are a more realistic approach. Our study shows that prompts constructed in this way do not perform as well as comprehensive narrative texts. We are not able to determine if this is due to failure to include some of the original information in the feature-based prompts or if it is due to the different textual style. However, we suggest that prompts with texts as sophisticated as those of the NEJM case reports are not realistic representations of what can be generated in typical clinical contexts because of privacy concerns and because it would be too time consuming for physicians to write similar clinical vignettes based on information available in EHRs or other resources. Therefore, we posit that the performance obtained by our feature-based approaches (PHENO-C and PHENO-R) is a better estimate of the performance that can be expected for typical clinical usage. We have additionally shown that the performance of GPT-4 differed substantially between two versions. The lack of stability of the algorithm, together with the black-box nature of GPT-4, raises important questions about the clinical utility of general purpose LLMs for clinical diagnostics. Future research and algorithmic development is needed to determine the optimal approach to leveraging LLMs for clinical diagnosis.

## Data Availability

All data used in our manuscript is taken from published New England Journal of Medicine case reports.

## Funding

NICHD: 5R01HD103805-03

NIH OD: 5R24OD011883-06

3U24TR002306-04S1

Director, Office of Science, Office of Basic Energy Sciences of the U.S. Department of Energy Contract No. DE-AC02-05CH11231

## References

1. Krishnan, R., Rajpurkar, P. & Topol, E. J. Self-supervised learning in medicine and healthcare. Nat Biomed Eng 6, 1346–1352 (2022).

2. Thirunavukarasu, A. J. et al. Large language models in medicine. Nat. Med. 29, 1930–1940 (2023).

3. Lee, J. et al. BioBERT: a pre-trained biomedical language representation model for biomedical text mining. Bioinformatics 36, 1234–1240 (2020).

4. Moor, M. et al. Foundation models for generalist medical artificial intelligence. Nature 616, 259–265 (2023).

5. Ouyang, L. et al. Training language models to follow instructions with human feedback. arXiv [cs.CL] (2022).

6. Nori, H., King, N., McKinney, S. M., Carignan, D. & Horvitz, E. Capabilities of GPT-4 on Medical Challenge Problems. arXiv [cs.CL] (2023).

7. Singhal, K. et al. Towards Expert-Level Medical Question Answering with Large Language Models. arXiv [cs.CL] (2023).

8. Lee, P., Bubeck, S. & Petro, J. Benefits, Limits, and Risks of GPT-4 as an AI Chatbot for Medicine. N. Engl. J. Med. 388, 1233–1239 (2023).

9. Hirosawa, T. et al. Diagnostic Accuracy of Differential-Diagnosis Lists Generated by Generative Pretrained Transformer 3 Chatbot for Clinical Vignettes with Common Chief Complaints: A Pilot Study. Int. J. Environ. Res. Public Health 20, (2023).

10. Rao, A. et al. Assessing the Utility of ChatGPT Throughout the Entire Clinical Workflow. medRxiv (2023) doi:10.1101/2023.02.21.23285886.

11. Mehnen, L., Gruarin, S., Vasileva, M. & Knapp, B. ChatGPT as a medical doctor? A diagnostic accuracy study on common and rare diseases. bioRxiv (2023) doi:10.1101/2023.04.20.23288859.

12. Suhag, A. et al. ChatGPT: a pioneering approach to complex prenatal differential diagnosis. Am J Obstet Gynecol MFM 5, 101029 (2023).

13. Koga, S., Martin, N. B. & Dickson, D. W. Evaluating the performance of large language models: ChatGPT and Google Bard in generating differential diagnoses in clinicopathological conferences of neurodegenerative disorders. Brain Pathol. e13207 (2023).

14. Kanjee, Z., Crowe, B. & Rodman, A. Accuracy of a Generative Artificial Intelligence Model in a Complex Diagnostic Challenge. JAMA (2023) doi:10.1001/jama.2023.8288.

15. Harry Caufield, J. et al. Structured prompt interrogation and recursive extraction of semantics (SPIRES): A method for populating knowledge bases using zero-shot learning. arXiv [cs.AI] (2023).

16. Köhler, S. et al. The Human Phenotype Ontology in 2021. Nucleic Acids Res. 49. D1207–D1217 (2021).

17. Reddy, K. P., Price, M. C., Barnes, J. A., Rigotti, N. A. & Crotty, R. K. Case 26-2022: A 48-Year-Old Woman with Cystic Lung Disease. N. Engl. J. Med. 387, 738–747 (2022).

18. Köhler, S. et al. Expansion of the Human Phenotype Ontology (HPO) knowledge base and resources. Nucleic Acids Res. 47. D1018–D1027 (2019).

19. Köhler, S. et al. The Human Phenotype Ontology project: linking molecular biology and disease through phenotype data. Nucleic Acids Res. 42. D966–D974 (2014).

20. Egli, A. ChatGPT, GPT-4, and other large language models - the next revolution for clinical microbiology? Clin. Infect. Dis. (2023) doi:10.1093/cid/ciad407.

21. Bender, E. M., Gebru, T., McMillan-Major, A. & Shmitchell, S. On the Dangers of Stochastic Parrots: Can Language Models Be Too Big? LJ. in Proceedings of the 2021 ACM Conference on Fairness, Accountability, and Transparency 610–623 (Association for Computing Machinery, 2021).

22. Zhang, X. A. et al. Semantic integration of clinical laboratory tests from electronic health records for deep phenotyping and biomarker discovery. NPJ Digit Med 2, (2019).

