## Supplement for "On the limitations of large language models in clinical diagnosis"

```
I am running an experiment on a clinicopathological case conference to
 see how your diagnoses compare with those of human experts. I am
going to give you part of a medical case. These have all been
published in the New England Journal of Medicine. You are not trying
to treat any patients. As you read the case, you will notice that
there are expert discussants giving their thoughts. In this case, you
are "Dr. GPT-4," an Al language model who is discussing the case along
 with human experts. A clinicopathological case conference has several
 unspoken rules. The first is that there is most often a single
definitive diagnosis (though rarely there may be more than one), and
it is a diagnosis that is known today to exist in humans. The
diagnosis is almost always confirmed by some sort of clinical
pathology test or anatomic pathology test, though in rare cases when
such a test does not exist for a diagnosis the diagnosis can instead
be made using validated clinical criteria or very rarely just
confirmed by expert opinion. You will be told at the end of the case
description whether a diagnostic test/tests are being ordered, which
you can assume will make the diagnosis/diagnoses. After you read the
case, I want you to give two pieces of information. The first piece of
 information is your most likely
diagnosis/diagnoses. You need to be as specific as possible -- the
goal is to get the correct answer, not a broad category of answers.
You do not need to explain your reasoning, just give the diagnosis/
diagnoses. The second piece of information is to give a robust
differential diagnosis, ranked by their probability so that the most
likely diagnosis is at the top, and the least likely is at the bottom.
 There is no limit to the number of diagnoses on your differential.
You can give
as many diagnoses as you think are reasonable. You do not need to
explain your reasoning, just list the diagnoses. Again, the goal is to
 be as specific as possible with each of the diagnoses.
Do you have any questions, Dr. GPT-4?

Here is the case:

...
```

**Figure S1**: **Prompt preamble**. Preamble for all GPT4 prompts used in this work. ”...” indicates that the specific content shown in Supplemental Figures XYZ is appended to the same preamble for all prompt construction methods.

```
Here is the case:

A 33-year-old pregnant woman with ulcerative colitis was admitted to
this hospital during the summer at 10 weeks of gestation because of
fever, nausea, vomiting, abdominal pain and tenderness, and headache.
The patient had been in her usual state of health until 3 days before
this admission, when fever, rigors, nausea, and vomiting developed.
During the next 3 days, the symptoms worsened; the patient was unable
to eat, drink, or take medications. She reported abdominal pain and
cramping on the left side that she described as being similar to
previous flares of ulcerative colitis; she also had nonbloody diarrhea
. The patient called the obstetrics clinic and was instructed to seek
evaluation in the emergency department of this hospital. A review of
systems was notable for fatigue, mild headache, neck pain, and
photophobia. The patient reported no hematochezia, pelvic pain,
vaginal bleeding, dysuria, or joint pain. Ulcerative colitis had been
diagnosed 11 years earlier; at the time of the diagnosis, she began
treatment with azathioprine but stopped after 1 year because of severe
 fatigue. The patient had taken mesalamine and sulfasalazine
intermittently for disease flares but typically stopped after a few
months of treatment. Exacerbations of ulcerative colitis occurred
approximately every year and were associated with fevers, rigors,
nausea, vomiting, abdominal pain, and bloody diarrhea. Nine months
before this admission, Clostridioides difficile colitis developed, and
 the patient was successfully treated with oral vancomycin. Three
weeks before the current admission, she was admitted to this hospital
for a flare of ulcerative colitis; intravenous methylprednisolone,
hydromorphone, and ondansetron were administered, and mesalamine was
restarted. Flexible sigmoidoscopy revealed mild erythema and friable
mucosa with a decreased vascular pattern in the sigmoid colon.
```

**Figure S2**: **Narrative Prompt**. In this example, the narrative text from the original New England Journal of Medicine (NEJM) case report: Ciaranello AL, et al. (2021) Case 21-2021: A 33-Year-Old Pregnant Woman with Fever, Abdominal Pain, and Headache. *N Engl J Med* **385**:265-274; PMID:34260840. The same example is used in Supplemetary Figures **S3**–

```
Here is the case:

A 33-year-old pregnant woman with ulcerative colitis was admitted to
this hospital during the summer at 10 weeks of gestation because of
fever, nausea, vomiting, abdominal pain and tenderness, and headache.
11 years earlier, the patient presented with Bloody diarrhea, Rigors,
Abdominal pain, Fatigue, Nausea and vomiting, Fever, and Ulcerative
colitis.
3 days before this admission, the patient presented with Rigors,
Nausea and vomiting, and Fever.
During the next 3 days, the patient presented with Abdominal pain,
Diarrhea, and Ulcerative colitis.
Nine months before this admission, the patient presented with Colitis.
Three weeks before the current admission, the patient presented with
Erythema, and Ulcerative colitis.
In the emergency department, the patient presented with Neck pain,
Headache, Photophobia, Fatigue, and Ulcerative colitis. The following
signs and symptoms were excluded: Arthralgia, Hematochezia, Pelvic
pain, and Dysuria.
```

**Figure S3**: **Phenotypic feature prompt generation - random (PHENO-R)**. Here, the text mining was used to extract Human Phenotype Ontology (HPO) terms from the narrative text shown in Supplementary Figure **S2**, and the prompt text was generated automatically by including the first sentence from the NEJM case report following by lists of clinical signs, symptoms, and findings, arranged according to time of presentation. In this method, the time periods are arranged randomly rather than chronologically.

```
Here is the case:


A 33-year-old pregnant woman with ulcerative colitis was admitted to
this hospital during the summer at 10 weeks of gestation because of
fever, nausea, vomiting, abdominal pain and tenderness, and headache.
3 days before this admission, the patient presented with Rigors,
Nausea and vomiting, and Fever.

During the next 3 days, the patient presented with Abdominal pain,
Diarrhea, and Ulcerative colitis.

In the emergency department, the patient presented with Neck pain,
Headache, Photophobia, Fatigue, and Ulcerative colitis.
The following signs and symptoms were excluded: Arthralgia,
Hematochezia, Pelvic pain, and Dysuria.
11 years earlier, the patient presented with Bloody diarrhea, Rigors,
Abdominal pain, Fatigue, Nausea and vomiting, Fever, and Ulcerative
colitis.

Nine months before this admission, the patient presented with Colitis.

Three weeks before the current admission, the patient presented with
Erythema and Ulcerative colitis.
```

**Figure S4**: **Phenotypic feature prompt generation - chronological (PHENO-C)**. This prompt is similar to the previous feature-based prompt (Supplemental Figure **S3**) but the various parts of the clinical source are arranged chronologically rather than randomly.

```
Here is the case:

A 33-year-old pregnant woman with ulcerative colitis was admitted to
this hospital during the summer at 10 weeks of gestation because of
fever, nausea, vomiting, abdominal pain and tenderness, and headache.
The past medical history was notable for Ulcerative colitis.
3 days before this admission, the patient presented with Rigors,
Nausea and vomiting, and Fever.
During the next 3 days, the patient presented with Abdominal pain,
Diarrhea, and Ulcerative colitis.
In the emergency department, the patient presented with Neck pain,
Headache, Photophobia, Fatigue, and Ulcerative colitis.
 The following signs and symptoms were excluded: Arthralgia,
 Hematochezia, Pelvic pain, and Dysuria.
11 years earlier, the patient presented with Bloody diarrhea, Rigors,
Abdominal pain, Fatigue, Nausea and vomiting, Fever, and Ulcerative
colitis.
 The following treatments were administered: mesalamine.
Nine months before this admission, the patient presented with Colitis
and Clostridioides difficile colitis.
Three weeks before the current admission, the patient presented with
Erythema and Ulcerative colitis.
 The following treatments were administered: mesalamine, hydromorphone
 , ondansetron, and intravenous methylprednisolone.
Flexible sigmoidoscopy revealed mild erythema and friable mucosa with
a decreased vascular pattern in the sigmoid colon
```

**Figure S5**: **Manual/HPO Prompt generation (MAN-HPO)**. Here, the text mining was used to extract Human Phenotype Ontology (HPO) terms from the narrative text shown in Supplementary Figure **S2**, and the prompt text was generated automatically by including the first sentence from the NEJM case report following by lists of clinical signs, symptoms, and findings, arranged according to time of presentation. In this method, the time periods are arranged randomly rather than chronologically.
